# Association of Long COVID with housing insecurity in the United States, 2022-2023

**DOI:** 10.1101/2023.06.05.23290930

**Authors:** Samuel E. Packard, Ezra Susser

**Affiliations:** Department of Epidemiology, Mailman School of Public Health, New York, NY; New York State Psychiatric Institute, New York, NY

**Keywords:** Housing, Housing Insecurity, Long COVID, Post-Acute COVID-19 Syndrome, COVID-19

## Abstract

**Objectives:** To assess the association of Long COVID with housing insecurity in the United States.

**Methods:** To compare the prevalence of 3 binary indicators of housing insecurity between people with Long COVID (symptoms > 3 months) and COVID-19 survivors who don’t report long-term symptoms, we used survey-weighted regression models on 203,807 responses from the Household Pulse Survey, a representative cross-sectional survey of US households collected September 2022 – April 2023. Among people with Long COVID, we assessed whether functional impairment, current COVID-19 related symptoms, and symptom impact on day-to- day life were associated with a higher prevalence of housing insecurity.

**Results:** During the study period, 54,446 (27.2%) respondents with COVID-19 experienced symptoms lasting 3 months or longer, representing an estimated 27 million US adults. People with Long COVID were nearly twice as likely to experience significant difficulty with household expenses (Prevalence ratio [PR] 1.85, 95% CI 1.74-1.96), be behind on housing payments (PR 1.76, 95% CI 1.57-1.99), and face likely eviction or foreclosure (PR 2.12, 95% CI 1.58-2.86). Functional limitation and current symptoms which impact day-to-day life were associated with higher prevalence of housing insecurity.

**Conclusions:** Compared with COVID-19 survivors who don’t experience long-term symptoms, people with Long COVID are more likely to report indicators housing insecurity, particularly those with functional limitations and long-term COVID-19 related symptoms impacting day-to- day life. Policies are needed to support people living with chronic illnesses following SARS-CoV-2 infection.

## 1. INTRODUCTION

Our lives are anchored around the places we call home. Finding and maintaining a safe, secure place of residence is among the foremost concerns for every person and family, intimately linked to nearly every aspect of our lives and well-being. At the population level, the determinants and consequences of access to housing are likewise fundamental to public health (Shaw, 2004). Not only do chronic health problems and disability adversely impact access to adequate and stable housing, but unstable housing is empirically related to subsequent physical and mental health outcomes (Cusack et al., 2021; Elder & King, 2019; Kang, 2022; Rao et al., 2023).

Unfortunately, many people in the United States face the very real threat of housing insecurity . The homeless population in the United States is over a half million people and growing, and prior to the COVID-19 pandemic more than 2 million households were being threatened with evictions in formal court filings each year, with evictions carried out on 6 - 7% of renting households (Gromis et al., 2022; National Alliance to End Homelessness, 2023). Early in the pandemic, housing insecurity was recognized as a significant public health issue likely to be exacerbated significantly the economic stresses experienced in 2020. As a result, policies were enacted at multiple levels of government, including a federal eviction moratorium issued by the Centers for Disease Control and Prevention, as US households struggled with the direct and indirect impacts of COVID-19 (Benfer et al., 2021; Raajkumar, 2022).

The end of the federal eviction moratorium in August 2021 coincided with an increasing awareness that a significant proportion of those who survive SARS-CoV-2 infection develop long-term sequelae, indicating a potential public health crisis affecting millions of people (Phillips & Williams, 2021). Long COVID, also known as Post-Acute Sequelae of COVID-19 and Post-COVID Conditions, is an umbrella term for a range of etiologies and illness experiences resulting from or triggered by COVID-19 infection, presenting with heterogeneous severity, duration, and symptomatology across multiple organ systems (Castanares-Zapatero et al., 2022; Davis et al., 2023; Michelen et al., 2021). Many people who develop Long COVID will experience profound livelihood changes due to the onset of significant physical or cognitive impairments, which have been linked to a number of adverse social and economic outcomes including social exclusion and reduced employment (Nittas et al., 2022). To date, however, no studies have assessed the degree to which people living with Long COVID face increased risk of housing insecurity.

Data from the Household Pulse Survey (HPS), a nationally representative survey of US households produced by the Census Bureau to monitor the social and economic impact of the COVID-19 pandemic(US Census Bureau, 2022c), have been used by researchers to describe the inter-relationship of housing insecurity with financial hardship, disability, and health status during various stages of the pandemic(Bushman & Mehdipanah, 2022; Friedman, 2023; Kim, 2021). In mid-2022, the survey added new questions about long term symptoms following acute COVID-19, making it the first source of nationally representative surveillance data on Long COVID in the United Sates (Levine, 2022; US Census Bureau, 2022b).

The present study uses these data to assess the association of Long COVID with housing insecurity among US adults with a history of COVID-19. Specifically, we hypothesized that compared to people who had COVID-19 but did not report long-term symptoms, those who report symptoms lasting longer than 3 months that they did not have prior to their infection (henceforth Long COVID) would be more likely to report indicators of housing insecurity including significant difficulty with routine household expenses, being behind on rent or mortgage payments, and perceived likelihood of eviction or foreclosure. Additionally, we hypothesized that among people with Long COVID, measures of functional impairment, current symptoms, and impact of symptoms on day-to-day life would be associated with increased prevalence of each indicator of housing insecurity.

## 2. MATERIALS AND METHODS

### 2.1 Study Population

Respondents were recruited from a stratified sample of domiciles in the US Census Bureau’s Master Address File, with one adult from each household being invited by phone or email to complete a 20-minute online questionnaire (US Census Bureau, 2022a). We accessed Public Use Files from Phases 3.6 - 3.8 and utilized data from weeks 49 - 56, representing 8 waves of data collected over 7 months from September 14, 2022 – April 10, 2023 (US Census Bureau, 2023). We excluded survey responses without confirmed history of COVID-19 infection (positive test result or diagnosis from healthcare provider) and those with onset of COVID-19 in the four weeks prior to taking the survey.

### 2.2 Measures

#### 2.2.1 Housing Tenure and Indicators of Housing Insecurity

Using data available from the HPS, we defined housing tenure as either owning a home or renting. A minimal proportion (< 1%) of non-homeowners reported occupying their home without making payments; these were respondents of predominantly low socio-economic status and collapsed into the “renting” category for this analysis. We operationalized three binary indicators of housing insecurity as outcome variables in this study. Significant difficulty with household expenses was defined as responding “Very difficult” to the question “In the last 7 days, how difficult has it been for your household to pay for usual household expenses, including but not limited to food, rent or mortgage, car payments, medical expenses, student loans, and so on?” Being behind on rent or mortgage payments was defined as responding “No” to the questions “Is the household currently caught up on [rent or mortgage] payments?” Perceived likelihood of eviction or foreclosure was defined as responding “Somewhat likely” or “Very likely” to the questions “How likely is it that your household will have to leave this home or apartment within the next two months because of [eviction or foreclosure]?” Respondents who reported not making rent or mortgage payments were considered not being behind on payments or at risk of eviction or foreclosure.

#### 2.2.2 Long COVID Status, Current Symptoms, and Measures of Functional Impairment

Those who reported ever testing positive or being told by a healthcare provider that they had COVID-19 were asked follow-up questions about long-term symptoms. We defined Long COVID as responding affirmatively to the question “Did you have any symptoms lasting 3 months or longer that you did not have prior to having coronavirus or COVID-19? Long term symptoms may include tiredness or fatigue, difficulty thinking, concentrating, forgetfulness, or memory problems (sometimes referred to as “brain fog”), difficulty breathing or shortness of breath, joint or muscle pain, fast-beating or pounding heart (also known as heart palpitations), chest pain, dizziness on standing, menstrual changes, changes to taste/smell, or inability to exercise.” We defined current symptoms as responding affirmatively to “Are you currently experiencing symptoms?”. Those who reported current symptoms were additionally asked “Do these long-term symptoms reduce your ability to carry out day-to-day activities compared with the time before you had COVID-19?” which we used to define symptom impact as “Not at all,” “A little,” or “A lot”.

Measures of functional impairment were collected on all survey respondents, unrelated to questions about COVID-19. Adapted from the Washington Group Short Set on Functioning (Weeks et al., 2021), these questions asked about difficulty with seeing, hearing, cognition, mobility, self-care, and communication. Within each domain, No Limitation was defined as responding “None at all”, Moderate Limitation was defined as responding “Some difficulty”, and Severe Limitation was defined as responding “A lot of difficulty” or “Cannot do at all”.

#### 2.2.3 Socio-Demographic Covariates

Demographic information collected on the survey included age, gender, educational attainment, race and ethnicity, marital status, and prior year household income. In this study, age and gender are hypothesized as confounders as they have been consistently associated with both the exposure and the outcome in previous studies (Davis et al., 2023; Michelen et al., 2021). While statistical adjustment for confounders can reduce bias in effect estimates, unnecessary adjustment for effect modifiers, colliders, and mediators can have the opposite effect of increasing bias (Wysocki et al., 2022). As race/ethnicity, income, education, and household size likely have a role in the hypothesized association but were not specifically hypothesized as confounders due to less consistent evidence in the Long COVID literature, they were added as covariates in a separate sensitivity analysis.

### 2.3 Statistical Analysis

#### 2.3.1 Descriptive Analysis

We used person-level survey weights provided by the HPS to estimate counts and proportions of study variables for the target population of US adults with a history of COVID-19. For bivariate analyses, we described the unadjusted survey-weighted proportions of housing insecurity indicators, socio-demographic covariates, functional limitations, and symptoms by Long COVID status.

#### 2.3.2 Statistical Models

To assess the association of housing insecurity and Long COVID, we used survey-weighted generalized linear models with a quasi-Poisson distribution, logarithmic link function, and robust standard errors to estimate adjusted prevalence ratios (Zou, 2004). Separate models were fitted for each indicator of housing insecurity in three sets of models as described below, adjusted for age, gender, and survey wave.

First, we regressed housing insecurity indicators on Long COVID, including an interaction term with housing tenure to assess whether the effect estimates differed between homeowners and renters. Second, we regressed indicators of housing insecurity on measures of functional limitation among people with Long COVID. Lastly, we regressed housing insecurity indicators on the presence or absence of current Long COVID symptoms and impact of these symptoms on day-to-day life among people with Long COVID. As a sensitivity analysis, we additionally adjusted each model for education, income, race/ethnicity, and household size. All analyses were performed in R version 4.2.2 using the survey package (Lumley, 2020; R Core Team, 2021). Conforming to Columbia University Institutional Review Boards policy on Use of Publicly Available Datasets for Research, formal review was not required.

## 3. RESULTS

### 3.1 Descriptive Analysis

A total of 203,807 survey respondents met inclusion criteria across the 8 waves of HPS data, representing an estimated 94,448,919 US adults with a history of confirmed SARS-CoV-2 infection, of whom an estimated 27,250,409 (28.8%) experienced new symptoms lasting 3 months or longer, meeting the study definition of Long COVID. One in five US adults with a history of COVID-19 (20.1%, 95% CI 19.7 – 20.1) experienced at least one measure of housing insecurity during the study period, including significant difficulty with household expenses (16.2%, 15.9 – 17.0), being behind on rent or mortgage payments (6.0%, 5.7 – 6.0), or being at risk of eviction or foreclosure (1.9%, 1.8 – 2.0). Inclusion in the study overall and for each sub-analysis is illustrated in Figure 1. Overall, 16% of eligible respondents were excluded due to missing data, and a comparison of included and excluded participants is provided in Table S1.

**Figure 1:**
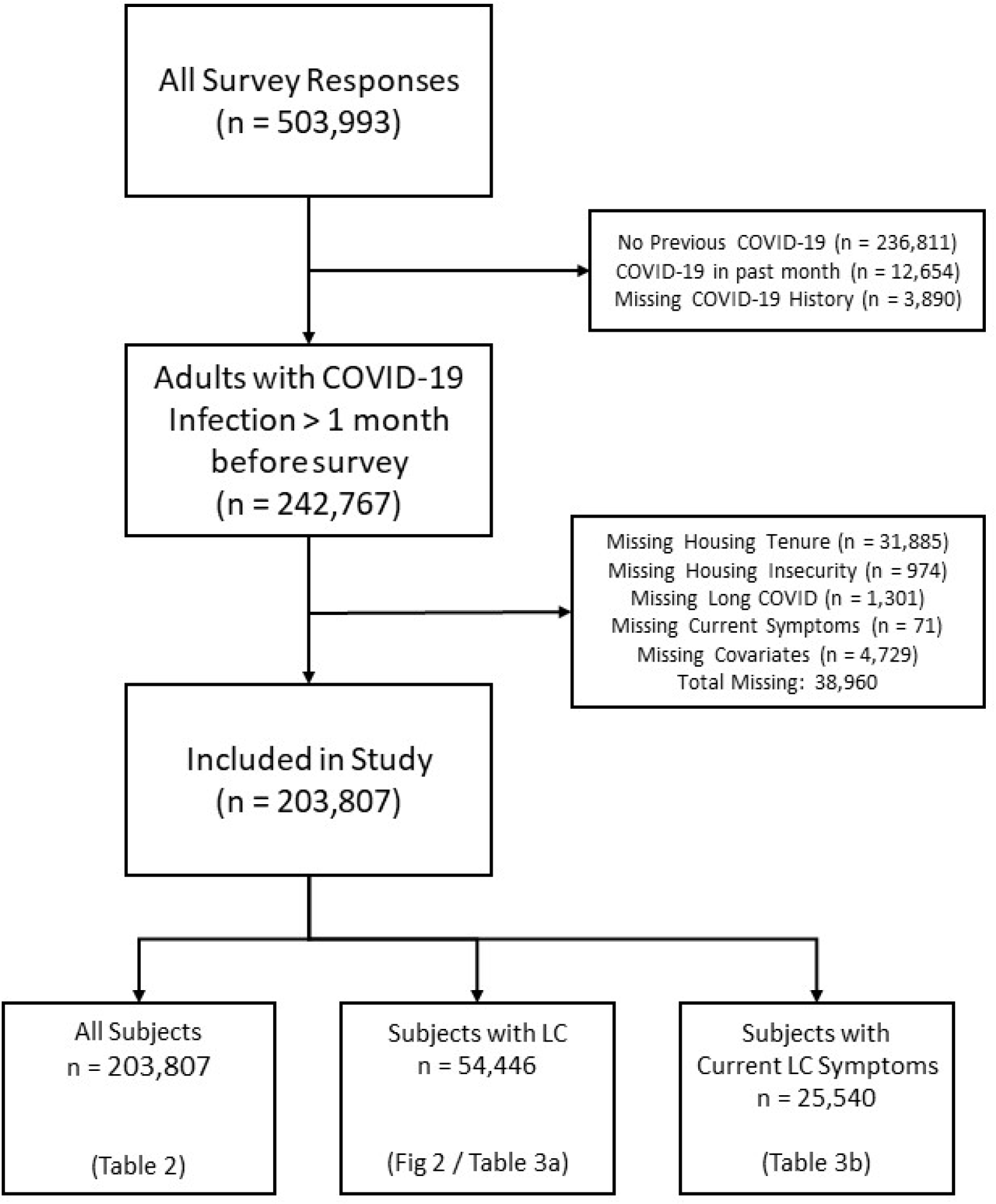
Inclusion of respondents in the overall study sample and each sub- analysis.

Demographic characteristics, housing data, and health measures are summarized by Long COVID status in Table 1. People with Long COVID were more likely than those without Long COVID to rent rather than own their home and more likely to report all three indicators of housing insecurity. People with Long COVID were also more likely to report moderate or severe functional limitation across each of the five domains assessed. For example, roughly two-thirds of people with Long COVID experienced cognitive limitations, nearly twice the prevalence of those without. Just under half of those with Long COVID were currently experiencing Long COVID symptoms at the time of the survey (43.8%, 43.0 – 45.0), and around 8 in 10 of these people reported that their symptoms reduced their ability to carry out day-to-day activities compared with the time preceding their active infection (79.8%, 78.9 – 81.0). People with Long COVID were additionally more likely to be female, transgender, or non-binary, more likely to be widowed, divorced, or separated, less likely to have a college degree, and more likely to report a 2021 household income of $50,000 or less.

**Table 1:** Selected demographic characteristics, indicators of housing insecurity, functional limitations, and COVID-19 symptoms by illness status among US adults with a history of COVID-19, September 2022 – April 2023.

| Characteristic | Long COVID | No Long COVID | Overall |
| --- | --- | --- | --- |
| Age |  |  |  |
| 18-24 | 1,891 (8.1%) | 4,845 (7.6%) | 6,736 (7.7%) |
| 25-44 | 22,292 (41.1%) | 61,473 (42.5%) | 83,765 (42.1%) |
| 45-64 | 22,538 (37.4%) | 52,620 (32.6%) | 75,158 (33.9%) |
| 65+ | 8,725 (13.4%) | 29,423 (17.3%) | 38,148 (16.2%) |
| Race/Ethnicity |  |  |  |
| White Non-Hispanic | 41,906 (64.0%) | 116,499 (66.8%) | 158,405 (66.0%) |
| White Hispanic | 4,589 (15.6%) | 9,791 (13.0%) | 14,380 (13.8%) |
| Black | 3,626 (9.1%) | 8,190 (8.7%) | 11,816 (8.8%) |
| Asian | 1,637 (3.8%) | 7,182 (5.9%) | 8,819 (5.3%) |
| Other | 3,688 (7.5%) | 6,699 (5.6%) | 10,387 (6.2%) |
| Gender |  |  |  |
| Male | 17,268 (36.1%) | 67,540 (50.1%) | 84,808 (46.1%) |
| Female | 36,800 (60.4%) | 78,506 (48.0%) | 115,306 (51.6%) |
| Transgender | 376 (NA%) | 488 (NA%) | 864 (0.7%) |
| None of the Above | 752 (1.9%) | 1,132 (0.9%) | 1,884 (1.2%) |
| Education |  |  |  |
| Less Than High School | 1,108 (6.5%) | 1,741 (4.6%) | 2,849 (5.1%) |
| High School Graduate | 6,565 (27.9%) | 13,367 (25.5%) | 19,932 (26.2%) |
| Some College or Associates | 21,099 (35.7%) | 39,938 (28.5%) | 61,037 (30.5%) |
| Bachelor's Degree | 14,965 (17.5%) | 48,074 (22.4%) | 63,039 (21.0%) |
| Graduate Degree | 11,709 (12.3%) | 45,241 (19.1%) | 56,950 (17.1%) |
| Income in 2021 |  |  |  |
| Less than \$25,000 | 6,453 (14.0%) | 8,614 (8.3%) | 15,067 (10.0%) |
| \$25,000 - \$34,999 | 5,269 (11.9%) | 8,794 (8.3%) | 14,063 (9.3%) |
| \$35,000 - \$49,999 | 6,565 (13.1%) | 12,553 (10.7%) | 19,118 (11.4%) |
| \$50,000 - \$74,999 | 9,902 (18.0%) | 22,153 (16.4%) | 32,055 (16.9%) |
| \$75,000 - \$99,999 | 7,924 (13.2%) | 21,075 (14.0%) | 28,999 (13.8%) |
| \$100,000 - \$149,999 | 9,551 (15.1%) | 30,471 (18.0%) | 40,022 (17.1%) |
| \$150,000 and above | 8,927 (12.8%) | 41,993 (22.1%) | 50,920 (19.4%) |
| Marital Status |  |  |  |
| Married | 30,398 (54.6%) | 94,109 (61.4%) | 124,507 (59.5%) |
| Widowed/Divorced/Separated | 13,121 (19.2%) | 24,909 (14.1%) | 38,030 (15.5%) |
| Never Married | 11,772 (25.8%) | 29,017 (24.3%) | 40,789 (24.7%) |
| Household Size | 3.4 (1.7) | 3.3 (1.6) | 3.3 (1.7) |
| Housing Tenure |  |  |  |
| Owned with mortgage/loan | 27,492 (46.9%) | 78,457 (49.8%) | 105,949 (49.0%) |
| Owned, no mortgage | 11,333 (19.5%) | 35,398 (22.6%) | 46,731 (21.7%) |
| Rented | 15,746 (31.9%) | 33,031 (26.4%) | 48,777 (28.0%) |
| Occupied without rent payments | 875 (1.7%) | 1,475 (1.2%) | 2,350 (1.4%) |
| Current on Rent/Mortgage Payments |  |  |  |
| Current on Rent/Mortgage | 39,447 (88.7%) | 106,886 (93.7%) | 146,333 (92.2%) |
| Behind on Payments | 3,791 (11.3%) | 4,602 (6.3%) | 8,393 (7.8%) |
| Months Behind on Rent/Mortgage Payments |  |  |  |
| Current | 39,447 (88.7%) | 106,886 (93.7%) | 146,333 (92.2%) |
| 0-1 Months Behind | 1,811 (5.3%) | 2,441 (3.0%) | 4,252 (3.7%) |
| 2-3 Months Behind | 1,186 (3.7%) | 1,190 (1.7%) | 2,376 (2.3%) |
| 4+ Months Behind | 707 (2.1%) | 712 (1.0%) | 1,419 (1.3%) |
| Eviction/Foreclosure in Next 2 Months |  |  |  |
| Very Likely | 421 (1.4%) | 352 (0.5%) | 773 (0.8%) |
| Somewhat Likely | 936 (2.9%) | 819 (1.2%) | 1,755 (1.7%) |
| Not Very Likely | 1,359 (3.9%) | 1,480 (2.1%) | 2,839 (2.7%) |
| Not Likely at All | 1,075 (3.1%) | 1,951 (2.4%) | 3,026 (2.6%) |
| Current on Payments | 39,447 (88.7%) | 106,886 (93.7%) | 146,333 (92.2%) |
| Difficulty Making Household Expenses |  |  |  |
| Not at all difficult | 14,501 (20.0%) | 69,074 (36.7%) | 83,575 (31.9%) |
| A little difficult | 16,975 (29.7%) | 42,169 (30.5%) | 59,144 (30.3%) |
| Somewhat difficult | 12,939 (26.0%) | 23,760 (19.8%) | 36,699 (21.6%) |
| Very difficult | 11,031 (24.3%) | 13,358 (12.9%) | 24,389 (16.2%) |
| Difficulty seeing |  |  |  |
| No limitation | 29,901 (52.6%) | 106,136 (69.7%) | 136,037 (64.8%) |
| Moderate limitation | 22,373 (41.0%) | 38,480 (27.1%) | 60,853 (31.1%) |
| Severe limitation | 3,103 (6.2%) | 3,557 (3.0%) | 6,660 (3.9%) |
| Difficulty hearing |  |  |  |
| No limitation | 41,455 (75.5%) | 122,309 (83.2%) | 163,764 (80.9%) |
| Moderate limitation | 11,735 (20.2%) | 22,605 (14.2%) | 34,340 (15.9%) |
| Severe limitation | 2,052 (4.0%) | 3,005 (2.3%) | 5,057 (2.8%) |
| Difficulty cognitive |  |  |  |
| No limitation | 19,251 (34.2%) | 94,064 (61.5%) | 113,315 (53.6%) |
| Moderate limitation | 28,779 (51.1%) | 48,248 (33.3%) | 77,027 (38.4%) |
| Severe limitation | 7,321 (14.5%) | 5,788 (5.0%) | 13,109 (7.8%) |
| Difficulty with mobility |  |  |  |
| No limitation | 35,937 (63.8%) | 123,370 (81.5%) | 159,307 (76.4%) |
| Moderate limitation | 15,046 (27.3%) | 20,688 (14.8%) | 35,734 (18.4%) |
| Severe limitation | 4,382 (8.6%) | 4,062 (3.5%) | 8,444 (5.0%) |
| Difficulty with self-care |  |  |  |
| No limitation | 49,023 (86.3%) | 141,798 (94.2%) | 190,821 (91.9%) |
| Moderate limitation | 5,434 (11.3%) | 5,587 (4.8%) | 11,021 (6.7%) |
| Severe limitation | 914 (2.2%) | 721 (0.7%) | 1,635 (1.2%) |
| Difficulty communicating |  |  |  |
| No limitation | 49,267 (86.9%) | 141,876 (94.1%) | 191,143 (92.1%) |
| Moderate limitation | 5,529 (11.4%) | 5,740 (5.0%) | 11,269 (6.9%) |
| Severe limitation | 589 (1.5%) | 555 (0.7%) | 1,144 (0.9%) |
| Currently experiencing COVID-19 symptoms |  |  |  |
| No | 29,898 (55.5%) | 141,055 (99.2%) | 170,953 (86.1%) |
| Yes | 25,548 (44.5%) | 1,149 (0.8%) | 26,697 (13.9%) |
| Long COVID Symptom impact on day-to-day activities |  |  |  |
| Not at all | 5,430 (20.2%) |  | 5,430 (20.2%) |
| A little | 14,597 (55.5%) |  | 14,597 (55.5%) |
| A lot | 5,513 (24.3%) |  | 5,513 (24.3%) |
| Note. Results presented as unweighted n (weighted %). |  |  |  |

### 3.2 Long COVID and Housing Insecurity

#### 3.2.1 Long COVID Status and Housing Insecurity

Models of the association of Long COVID with housing insecurity and its interaction with housing tenure are summarized in Table 2. The adjusted prevalence of housing insecurity among people with Long COVID was nearly twice as high as those without Long COVID, measured as difficulty with household expenses (Prevalence ratio [PR] 1.82, CI 1.72 - 1.93; P<.001), being behind on rent or mortgage (PR 1.76, CI 1.57 - 1.98; P<.001), or facing likely eviction or foreclosure (PR 2.12, CI 1.58 - 2.85; P<.001). Households renting their homes were also 2 - 6 times more likely to report housing insecurity than homeowners. There was not consistent evidence for multiplicative interaction between these exposures, though Long COVID was slightly less associated with difficulty making household expenses for renters compared to homeowners.

**Table 2:**
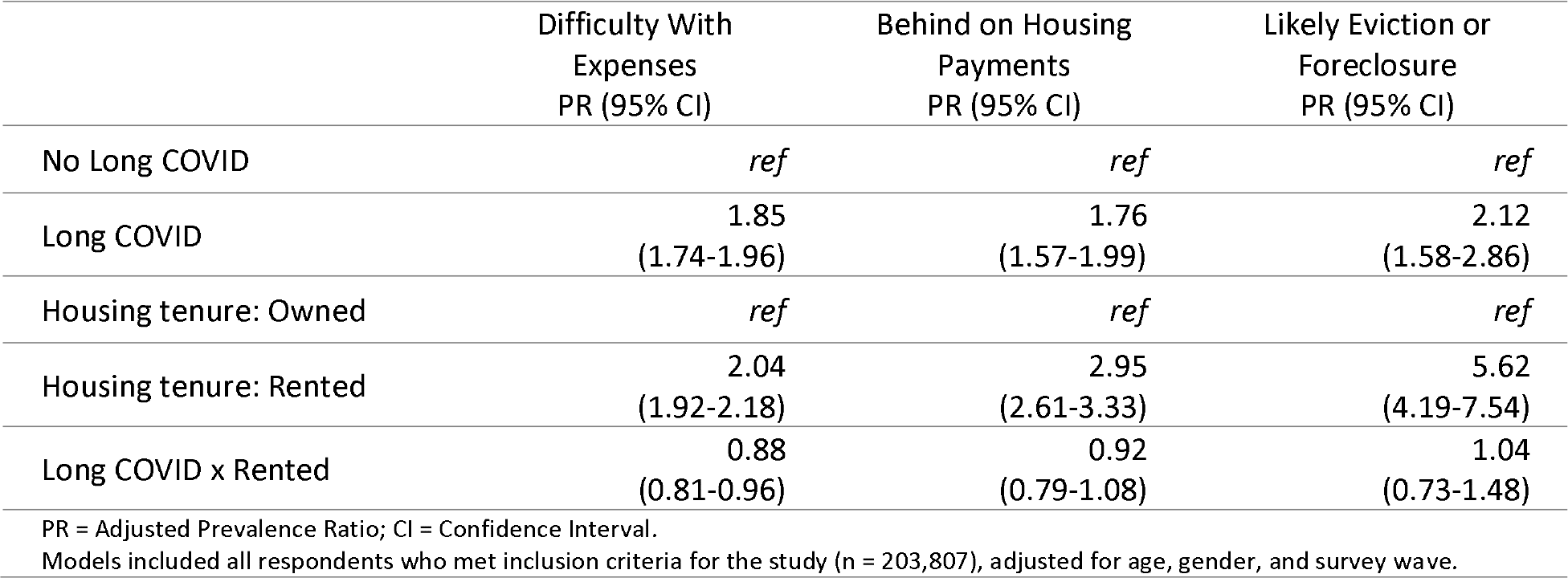
Association between Long COVID, Housing Tenure, and Housing Insecurity among adults with history of confirmed COVID-19 in the United States, September 2022 – April 2023.

|  | Difficulty With Expenses<br>PR (95% CI) | Behind on Housing Payments<br>PR (95% CI) | Likely Eviction or Foreclosure<br>PR (95% CI) |
| --- | --- | --- | --- |
| No Long COVID | *ref* | *ref* | *ref* |
| Long COVID | 1.85<br>(1.74-1.96) | 1.76<br>(1.57-1.99) | 2.12<br>(1.58-2.86) |
| Housing tenure: Owned | *ref* | *ref* | *ref* |
| Housing tenure: Rented | 2.04<br>(1.92-2.18) | 2.95<br>(2.61-3.33) | 5.62<br>(4.19-7.54) |
| Long COVID x Rented | 0.88<br>(0.81-0.96) | 0.92<br>(0.79-1.08) | 1.04<br>(0.73-1.48) |

PR = Adjusted Prevalence Ratio; CI = Confidence Interval. Models included all respondents who met inclusion criteria for the study (n = 203,807), adjusted for age, gender, and survey wave.

#### 3.2.2 Functional Impairment among People with Long COVID

Models of the association between functional impairment and housing insecurity among people with Long COVID and are summarized in Figure 2 and Table S2. There was a consistent trend of increased housing insecurity among people with Long COVID who reported moderate or severe functional limitation across all domains. Difficulty making household expenses was more strongly associated with severe limitation compared to moderate limitation, but this difference was less apparent for being behind on housing payments or being at risk of eviction or foreclosure.

**Figure 2:**
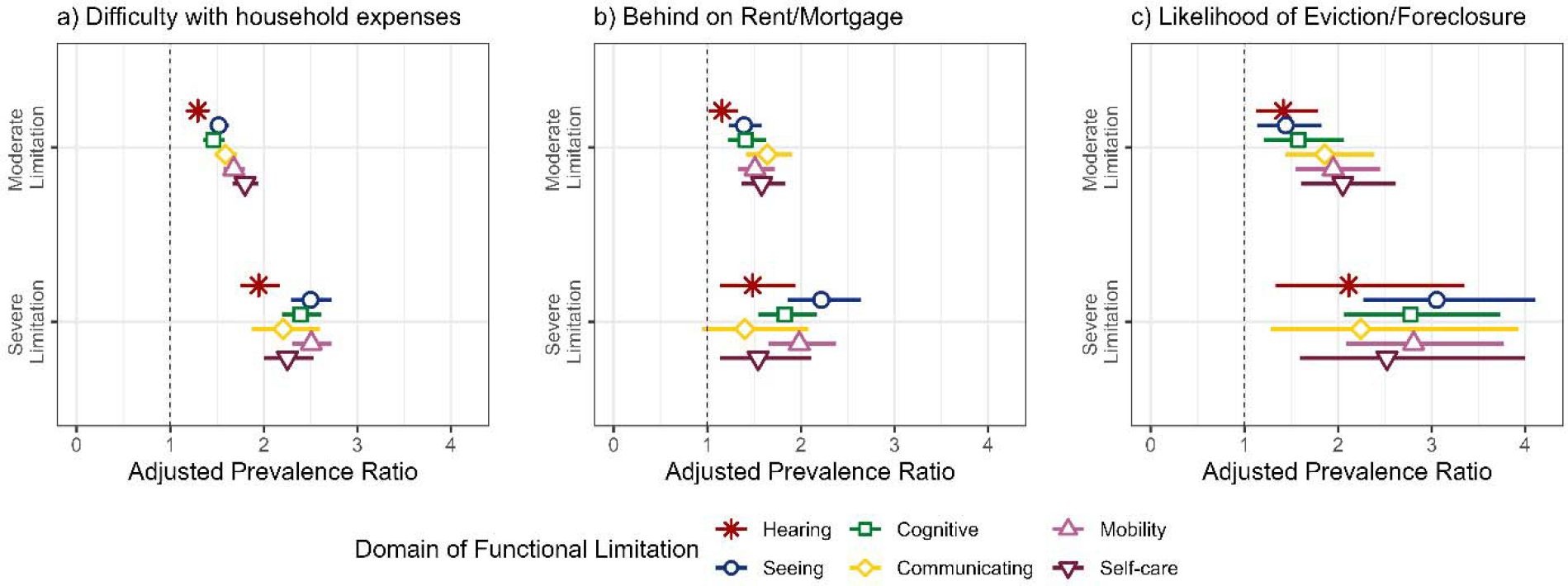
Association of Functional Limitations with Housing Insecurity Among Adults with Long COVID in the United States, September 2022 – April 2023. Estimates are displayed as Prevalence Ratios (PR) and 95% Confidence Intervals (CI), adjusted for age, gender, and survey wave. The reference group for each PR is respondents who reported no functional limitation. Models included all survey respondents with Long COVID (n = 54,446)

#### 3.2.3 Current COVID-related symptoms and symptom impact among People with Long COVID

Models estimating the association of currents symptoms and symptom impact with housing insecurity among people with Long COVID are summarized in Table 3. The adjusted prevalence of housing insecurity was slightly elevated among people whose symptoms were current compared to those not currently experiencing symptoms. Among those who reported current symptoms, the prevalence of housing insecurity was strongly associated with reported impact of symptoms on day-to-day life compared to the time preceding SARS-CoV-2 infection. Compared to those who reported that their current symptoms did not impact their day-to-day life compared to the time before they had COVID-19, those who reported that their symptoms impacted their life “a lot” were over 3 times more likely to report difficulty making household expenses (PR 3.41, CI 2.97 – 3.92) and being behind on rent (PR 3.50, CI 2.66 – 4.61), and nearly tenfold more likely to be at risk for eviction or foreclosure (PR 9.21, CI 5.55-15.29).

**Table 3:** Association between Symptoms and Housing Insecurity Among Adults with Long COVID in the United States, September 2022 – April 2023

|  | Difficulty With Expenses<br>PR (95% CI) | Behind on Housing Payments<br>PR (95% CI) | Likely Eviction or Foreclosure<br>PR (95% CI) |
| --- | --- | --- | --- |
| Symptoms: Not Current | *ref* | *ref* | *ref* |
| Symptoms: Current | 1.20<br>(1.13-1.28) | 1.09<br>(0.97-1.22) | 1.31<br>(1.06-1.62) |
| Symptom Impact: None | *ref* | *ref* | *ref* |
| Symptom Impact: A little | 1.73<br>(1.50-1.99) | 2.00<br>(1.52-2.63) | 3.92<br>(2.28-6.74) |
| Symptom Impact: A lot | 3.43<br>(2.98-3.94) | 3.54<br>(2.69-4.68) | 9.38<br>(5.59-15.75) |

PR = Adjusted Prevalence Ratio; CI = Confidence Interval. All models adjusted for age, gender, and survey wave. Models of current symptoms included all respondents with Long COVID (n = 54,446) Models of symptom impact included respondents who reported current symptoms (n = 25,540).

Sensitivity analyses are summarized in Tables S3 –S5. Additional adjustment in the models for education, income, race/ethnicity, and household size resulted in a small (∼10%) attenuations to the magnitudes of the effect sizes reported.

## 4. DISCUSSION

Based on a large, representative survey of US households during the third year of the COVID-19 pandemic, over 25 million US adults were estimated to have experienced long-term symptoms after infection with SARS-CoV-2. This represents 29% of COVID-19 survivors, which is higher than some conservative estimates of the prevalence of Long COVID but consistent with prior studies in the United States(O’Mahoney et al., 2023; Perlis et al., 2022; Robertson et al., 2023; Spatz et al., 2023). One third of people with Long COVID were either struggling to make household expenses, behind on housing payments, or facing likely eviction or foreclosure in the near future. These measures of housing insecurity were nearly twice as prevalent among people with Long COVID compared to those who had COVID-19 without long term symptoms, and significantly more prevalent among renters compared to homeowners. Among people living with Long COVID, housing insecurity was significantly elevated among those who reported that these long-term symptoms reduce their ability to carry out day-to-day activities compared to the time prior to their illness, and significantly elevated among those who specifically report moderate or severe functional impairments, particularly cognitive, mobility, and self-care limitations.

This research helps illuminate the profound ongoing social and economic consequences of the COVID-19 pandemic. Despite a significant decline in incident COVID-19 cases, hospitalizations, and deaths, the pandemic continues to impact millions of people in the US on a daily basis, particularly those survivors of SARS-CoV-2 infection who continue to experience long-term symptoms. While we are still learning about the underlying causes of Long COVID, it is clear that many survivors of COVID-19 experience lasting health problems, posing a significant population health challenge. Even after the end of the federal Public Health Emergency in the United Statues (Assistant Secretary for Public Affairs, US Department of Health and Human Services, 2023), is essential that sustained pandemic recovery efforts acknowledge and address the ongoing public health crisis of Long COVID and provide support through continued COVID-19 prevention, disability accommodations at work, and housing support and renter protection. As the manifold health and social consequences of housing insecurity and homelessness are well documented, addressing housing insecurity must be at the forefront of any effective effort to address Long COVID as public health crisis. The large and increasing number of people who are already homeless in the United States adds to the urgency.

As the directionality of the observed association was not established due to the cross-sectional study design, these results are open to various potential interpretations which inform the advancement of multiple hypotheses for future research. It is, for example, very plausible that the symptoms and functional limitations experienced by people with Long COVID result in disrupted livelihoods, reduced employment, or increased healthcare spending which effect subsequent challenges in being able to make routine household expenses, including rent or mortgage payments. Alternatively, it is also plausible that people with a history of unstable housing due to pre-existing socioeconomic disadvantage, disability, or chronic illness may be more prone to develop sequelae from COVID-19 due to material deprivation, stress, or other factors which could predispose someone to chronic illness triggered by viral infection. A better understanding of how the social and structural determinants of health shape the increased risk of socio-economic hardship faced by people with Long COVID should be a priority for future research, but regardless of the specific causal pathways involved, this evidence demonstrates that many people living with Long COVID face the added burden of social and economic precarity. Whether Long COVID precedes housing insecurity or vice-versa, it is important that policymakers, health providers, and public health officials are aware that a significant population of people who have not recovered from COVID-19 are struggling to make ends meet while effective treatments have yet to be developed.

Notably, the prevalence of both Long COVID and housing insecurity were elevated among respondents of low income and educational attainment, and adjustment for race, ethnicity, income, and education slightly attenuated the magnitude of the association between Long COVID and housing insecurity.

The strengths of this analysis include leveraging hundreds of thousands of survey responses over a 7-month period from a nationally representative survey during the third year of the COVID-19 pandemic, providing generalizable estimates of the downstream public health manifestations of the first several waves of infections. Despite these strengths, the following limitations should be considered when interpreting the results. First, the assessment of Long COVID status in the Household Pulse Survey was subject to several constraints. Only those who reported a positive COVID-19 test or physician confirmed diagnosis were asked about long-term symptoms, a conditional case definition for Long COVID which excludes people who were unable to access testing or healthcare at the time of their active infection for any reason, despite the fact that the social and structural drivers of inequality in access to these resources have been well-documented (Bilal et al., 2021; Mody et al., 2021). Determination of illness duration was also not possible from these data, and questions on the impact of Long COVID symptoms on day-to-day life were limited to only those experiencing symptoms at the time of the survey, despite previous research on episodic and relapsing patterns of symptoms in many people with Long COVID (Davis et al., 2021, 2023). Second, the potential for recall and selection bias may challenge the internal and external validity of our results. Recall bias, or any tendency of those experiencing housing insecurity to be systematically more or less likely to recall long term symptoms or attribute non-specific symptoms to COVID-19, could bias estimates away from or towards the null, respectively. Overall response rates to the survey are also low and participation from minoritized communities was generally lower than from non-Hispanic white residents. While it is important to consider that the analytic sample may not be completely representative, these threats to validity are mitigated by the sampling design and person-level weights based on the Census Bureau’s master address file which has been demonstrated to mitigate non-response bias in earlier iterations of the survey(Peterson et al., 2021).

In conclusion, this study draws attention to the fact that millions of people with Long COVID struggle with routine household expenses, including housing payments, and may be at risk of eviction or foreclosure. While longitudinal and qualitative data are needed to describe in more granular and causal terms how the illness experience of Long COVID relates to social and economic hardships in the context of other disruptive long-term impacts of the pandemic, it is clear that the pandemic continues to have deep and sustained impacts on those with viral onset chronic illness, and a comprehensive response and recovery effort will require acknowledging, monitoring, and supporting people with Long COVID.

## Disclosures

This work was supported by National Institutes of Mental Health [Grant number T32- MH013043, 2022]. The authors declare no conflicts of interest.

## Data Availability

Data used in this present work are publicly available online at https://www.census.gov/programs-surveys/household-pulse-survey/datasets.html

## ACKNOWLEDGEMENTS

We would like to thank Lisa McCorkell and Letícia Soares from the Patient-Led Research Collaborative for their helpful comments on a previous draft of this manuscript.

## Notes

### Competing Interest Statement

The authors have declared no competing interest.

### Funding Statement

This work was supported by National Institute of Mental Health #T32-MH013043

### Author Declarations

This study exclusively used publicly available, de-identified data from the US Census Bureau Household Pulse Survey. These data were made publicly available prior to the analyses performed in this study, and can be accessed at https://www.census.gov/programs-surveys/household-pulse-survey/datasets.html

### Summary of Updates

Added language in introduction section, more clarity was added in the discussion regarding interpretation of results.

