## Supplement for "Association of Long COVID with housing insecurity in the United States, 2022-2023"

Table S1: Comparison of respondents included in the study sample with eligible participants excluded due to missing data

| **Characteristic** | **Excluded**  N = 38,960*^1^* | **Included**  N = 203,807*^1^* | **Overall**  N = 242,767*^1^* | **p-value***^2^* |
| --- | --- | --- | --- | --- |
| Age | 43.0 (33.0, 57.0) | 47.0 (36.0, 61.0) | 47.0 (36.0, 60.0) | <0.001 |
| Race/Ethnicity |  |  |  | <0.001 |
| White Non-Hispanic | 26,207 (67.3%) | 158,405 (77.7%) | 184,612 (76.0%) |  |
| White Hispanic | 4,135 (10.6%) | 14,380 (7.1%) | 18,515 (7.6%) |  |
| Black | 4,172 (10.7%) | 11,816 (5.8%) | 15,988 (6.6%) |  |
| Asian | 2,065 (5.3%) | 8,819 (4.3%) | 10,884 (4.5%) |  |
| Other | 2,381 (6.1%) | 10,387 (5.1%) | 12,768 (5.3%) |  |
| Gender |  |  |  | <0.001 |
| Male | 15,677 (40.2%) | 84,808 (41.6%) | 100,485 (41.4%) |  |
| Female | 22,291 (57.2%) | 115,306 (56.6%) | 137,597 (56.7%) |  |
| Transgender | 143 (0.4%) | 864 (0.4%) | 1,007 (0.4%) |  |
| None of the Above | 474 (1.2%) | 1,884 (0.9%) | 2,358 (1.0%) |  |
| No Response | 375 (1.0%) | 945 (0.5%) | 1,320 (0.5%) |  |
| Education |  |  |  | <0.001 |
| Less Than High School | 1,246 (3.2%) | 2,849 (1.4%) | 4,095 (1.7%) |  |
| High School Graduate | 6,087 (15.6%) | 19,932 (9.8%) | 26,019 (10.7%) |  |
| Some College or Associates | 14,041 (36.0%) | 61,037 (29.9%) | 75,078 (30.9%) |  |
| Bachelors Degree | 10,275 (26.4%) | 63,039 (30.9%) | 73,314 (30.2%) |  |
| Graduate Degree | 7,311 (18.8%) | 56,950 (27.9%) | 64,261 (26.5%) |  |
| Income (2021) |  |  |  | <0.001 |
| Less than $25,000 | 230 (13.1%) | 15,067 (7.4%) | 15,297 (7.4%) |  |
| $25,000 - $34,999 | 110 (6.3%) | 14,063 (6.9%) | 14,173 (6.9%) |  |
| $35,000 - $49,999 | 148 (8.4%) | 19,118 (9.4%) | 19,266 (9.4%) |  |
| $50,000 - $74,999 | 169 (9.6%) | 32,055 (15.7%) | 32,224 (15.7%) |  |
| $75,000 - $99,999 | 136 (7.8%) | 28,999 (14.2%) | 29,135 (14.2%) |  |
| $100,000 - $149,999 | 142 (8.1%) | 40,022 (19.6%) | 40,164 (19.5%) |  |
| $150,000 and above | 175 (10.0%) | 50,920 (25.0%) | 51,095 (24.9%) |  |
| No Response | 643 (36.7%) | 3,563 (1.7%) | 4,206 (2.0%) |  |
| Marital Status |  |  |  | <0.001 |
| Married | 21,312 (54.7%) | 124,507 (61.1%) | 145,819 (60.1%) |  |
| Widowed/Divorced/Separated | 7,472 (19.2%) | 38,030 (18.7%) | 45,502 (18.7%) |  |
| Never Married | 9,985 (25.6%) | 40,789 (20.0%) | 50,774 (20.9%) |  |
| No Response | 191 (0.5%) | 481 (0.2%) | 672 (0.3%) |  |
| People in Household | 3.0 (2.0, 4.0) | 2.0 (2.0, 4.0) | 2.0 (2.0, 4.0) | <0.001 |
| Difficuly seeing |  |  |  | <0.001 |
| No difficulty | 4,446 (58.8%) | 136,037 (66.7%) | 140,483 (66.5%) |  |
| Moderate difficulty | 2,176 (28.8%) | 60,853 (29.9%) | 63,029 (29.8%) |  |
| Severe difficulty | 339 (4.5%) | 6,660 (3.3%) | 6,999 (3.3%) |  |
| Difficulty hearing |  |  |  | <0.001 |
| No difficulty | 5,543 (73.4%) | 163,764 (80.4%) | 169,307 (80.1%) |  |
| Moderate difficulty | 1,100 (14.6%) | 34,340 (16.8%) | 35,440 (16.8%) |  |
| Severe difficulty | 248 (3.3%) | 5,057 (2.5%) | 5,305 (2.5%) |  |
| Difficulty cognitive |  |  |  | <0.001 |
| No difficulty | 3,706 (49.1%) | 113,315 (55.6%) | 117,021 (55.4%) |  |
| Moderate difficulty | 2,604 (34.5%) | 77,027 (37.8%) | 79,631 (37.7%) |  |
| Severe difficulty | 567 (7.5%) | 13,109 (6.4%) | 13,676 (6.5%) |  |
| Difficulty with mobility |  |  |  | <0.001 |
| No difficulty | 5,185 (68.6%) | 159,307 (78.2%) | 164,492 (77.8%) |  |
| Moderate difficulty | 1,293 (17.1%) | 35,734 (17.5%) | 37,027 (17.5%) |  |
| Severe difficulty | 391 (5.2%) | 8,444 (4.1%) | 8,835 (4.2%) |  |
| Difficulty with self-care |  |  |  | <0.001 |
| No difficulty | 6,283 (83.2%) | 190,821 (93.6%) | 197,104 (93.3%) |  |
| Moderate difficulty | 473 (6.3%) | 11,021 (5.4%) | 11,494 (5.4%) |  |
| Severe difficulty | 104 (1.4%) | 1,635 (0.8%) | 1,739 (0.8%) |  |
| Difficulty communicating |  |  |  | <0.001 |
| No difficulty | 6,251 (82.7%) | 191,143 (93.8%) | 197,394 (93.4%) |  |
| Moderate difficulty | 521 (6.9%) | 11,269 (5.5%) | 11,790 (5.6%) |  |
| Severe difficulty | 86 (1.1%) | 1,144 (0.6%) | 1,230 (0.6%) |  |
| Current symptoms |  |  |  | <0.001 |
| 0 | 31,623 (88.5%) | 170,953 (86.5%) | 202,576 (86.8%) |  |
| 1 | 4,100 (11.5%) | 26,697 (13.5%) | 30,797 (13.2%) |  |
| Symptom impact on day-to-day activities |  |  |  | 0.033 |
| Not at all | 807 (20.9%) | 5,430 (21.3%) | 6,237 (21.2%) |  |
| A little | 2,146 (55.6%) | 14,597 (57.2%) | 16,743 (57.0%) |  |
| A lot | 904 (23.4%) | 5,513 (21.6%) | 6,417 (21.8%) |  |
| *^1^* Median (IQR); n (%) *^2^* Welch Two Sample t-test; Pearson's Chi-squared test | | | | |

Table S2: Associations of Functional Limitations with Housing Insecurity Among Adults with Long COVID in the United States, September 2022 – April 2023.

| Housing Insecurity Indicator | Domain of Limitation | Moderate Limitation  PR [95% CI] | Severe Limitation  PR [95% CI] |
| --- | --- | --- | --- |
| Difficulty with Household Expenses | hearing | 1.30 *** [1.20, 1.39] | 1.95 *** [1.75, 2.17] |
|  | seeing | 1.52 *** [1.42, 1.63] | 2.50 *** [2.29, 2.73] |
|  | remembering | 1.46 *** [1.35, 1.58] | 2.39 *** [2.19, 2.61] |
|  | understand | 1.59 *** [1.47, 1.72] | 2.21 *** [1.88, 2.60] |
|  | mobility | 1.68 *** [1.57, 1.80] | 2.51 *** [2.31, 2.73] |
|  | selfcare | 1.80 *** [1.67, 1.94] | 2.25 *** [2.01, 2.53] |
| Behind on Rent/Mortgage | hearing | 1.16 * [1.01, 1.33] | 1.48 ** [1.13, 1.95] |
|  | seeing | 1.39 *** [1.23, 1.58] | 2.22 *** [1.86, 2.65] |
|  | remembering | 1.41 *** [1.22, 1.63] | 1.83 *** [1.54, 2.17] |
|  | understand | 1.64 *** [1.41, 1.91] | 1.40 [0.94, 2.08] |
|  | mobility | 1.51 *** [1.33, 1.72] | 1.98 *** [1.66, 2.37] |
|  | selfcare | 1.58 *** [1.36, 1.83] | 1.54 ** [1.13, 2.11] |
| Likely Eviction/Foreclosure | hearing | 1.42 ** [1.12, 1.78] | 2.12 ** [1.33, 3.35] |
|  | seeing | 1.44 ** [1.14, 1.83] | 3.05 *** [2.27, 4.11] |
|  | remembering | 1.58 *** [1.21, 2.06] | 2.77 *** [2.06, 3.73] |
|  | understand | 1.86 *** [1.44, 2.39] | 2.25 ** [1.29, 3.93] |
|  | mobility | 1.95 *** [1.55, 2.45] | 2.81 *** [2.09, 3.77] |
|  | selfcare | 2.05 *** [1.61, 2.61] | 2.52 *** [1.59, 4.00] |

Table S3: Association between Long COVID, Housing Tenure, and Housing Insecurity among adults with history of confirmed COVID-19 in the United States, September 2022 – April 2023. Adjusted for age, gender, survey wave, race/ethnicity, income, education, and household size.

|  | Difficulty With Expenses | Behind on Rent/Mortgage | Likely Eviction/Foreclosure |
| --- | --- | --- | --- |
| Long COVID | 1.66 ***  (1.57-1.76) | 1.60 ***  (1.42-1.80) | 1.85 ***  (1.37-2.50) |
| Rented | 1.87 ***  (1.76-1.99) | 2.76 ***  (2.45-3.11) | 4.86 ***  (3.63-6.50) |
| Long COVID x Rented | 0.88 **  (0.81-0.96) | 0.92  (0.78-1.08) | 1.05  (0.74-1.49) |
| *** p < 0.001; ** p < 0.01; * p < 0.05. | | | |

Table S4: Association between Symptoms and Housing Insecurity Among Adults with Long COVID in the United States, September 2022 – April 2023. Adjusted for age, gender, survey wave, race/ethnicity, income, education, and household size.

|  | Difficulty w/ Expenses | Behind on Payments | Likely Eviction/  Foreclosure |
| --- | --- | --- | --- |
| Current Symptoms | 1.23 ***  (1.16-1.30) | 1.14 *  (1.01-1.27) | 1.39 **  (1.13-1.72) |
| Symptom Impact - A little | 1.68 ***  (1.46-1.93) | 1.92 ***  (1.46-2.52) | 3.60 ***  (2.12-6.12) |
| Symptom Impact - A lot | 3.06 ***  (2.66-3.52) | 3.04 ***  (2.29-4.04) | 7.53 ***  (4.45-12.74) |

Table S5: Associations of Functional Limitations with Housing Insecurity Among Adults with Long COVID in the United States, September 2022 – April 2023. Adjusted for age, gender, survey wave, race/ethnicity, income, education, and household size.

| Housing Insecurity Indicator | Domain of Limitation | Moderate Limitation  PR [95% CI] | Severe Limitation  PR [95% CI] |
| --- | --- | --- | --- |
| Difficulty with Household Expenses | hearing | 1.30 *** [1.21, 1.39] | 1.80 *** [1.61, 2.02] |
|  | seeing | 1.45 *** [1.36, 1.55] | 2.24 *** [2.05, 2.45] |
|  | remembering | 1.47 *** [1.36, 1.59] | 2.30 *** [2.12, 2.51] |
|  | understand | 1.52 *** [1.40, 1.64] | 1.89 *** [1.60, 2.24] |
|  | mobility | 1.59 *** [1.49, 1.70] | 2.22 *** [2.04, 2.42] |
|  | selfcare | 1.70 *** [1.58, 1.82] | 2.01 *** [1.77, 2.29] |
| Behind on Rent/Mortgage | hearing | 1.23 ** [1.08, 1.41] | 1.42 * [1.09, 1.86] |
|  | seeing | 1.35 *** [1.20, 1.53] | 1.99 *** [1.67, 2.38] |
|  | remembering | 1.49 *** [1.29, 1.71] | 1.90 *** [1.60, 2.24] |
|  | understand | 1.59 *** [1.37, 1.84] | 1.20 [0.80, 1.80] |
|  | mobility | 1.45 *** [1.27, 1.64] | 1.77 *** [1.48, 2.13] |
|  | selfcare | 1.51 *** [1.30, 1.75] | 1.40 * [1.02, 1.92] |
| Likely Eviction/Foreclosure | hearing | 1.55 *** [1.24, 1.94] | 2.00 ** [1.27, 3.14] |
|  | seeing | 1.39 ** [1.10, 1.75] | 2.66 *** [1.97, 3.58] |
|  | remembering | 1.72 *** [1.32, 2.25] | 2.93 *** [2.17, 3.96] |
|  | understand | 1.78 *** [1.40, 2.28] | 1.80 * [1.00, 3.23] |
|  | mobility | 1.87 *** [1.49, 2.35] | 2.45 *** [1.80, 3.33] |
|  | selfcare | 1.96 *** [1.54, 2.49] | 2.24 *** [1.41, 3.55] |
